# Prevalence And Factors Associated With Surgical Site Infection In Patients Undergoing Hepatopancreaticobiliary Surgery, a Prospective Multi-Centric Cross-Sectional Study in Addis Ababa, Ethiopia

**DOI:** 10.64898/2025.12.28.25343114

**Authors:** Gemeda Nebi Edasu, Abigael Abiy Mesfin, Samuel Zerihun Tesfaye, Sahlu Wondimu

**Affiliations:** Hepatopancreaticobiliary Unit, Department of Surgery, School of Medicine, College of Health Sciences, Addis Ababa University, Addis Ababa, Ethiopia; School of Medicine, College of Health Sciences, Addis Ababa University, Addis Ababa, Ethiopia

**Keywords:** surgical, site, infections, Ethiopia, hepatobiliary

## Abstract

**Background:** Surgical site infections are potentially preventable complications following surgery and impose a significant burden in terms of patient morbidity, mortality, increased cost of treatment, and diminished quality of life for patients. The prevalence of SSIs in hepatopancreatobiliary surgery varies widely across different studies and settings. Therefore, this study aimed to determine the prevalence and risk factors associated with SSIs in HPB surgery in selected hospitals in Addis Ababa, Ethiopia.

**Methods:** This hospital-based prospective cross-sectional study was conducted in three major hospitals in Addis Ababa, Ethiopia: Tikur Anbessa Specialized Hospital, Yekatit 12 Specialized Hospital, and Menelik II Specialized Hospital. The study took place over a period of 6 months. It commenced on January 10, 2025 and concluded on June 30 2025. Bivariate and multivariate logistic regression analyses were performed to evaluate the associations of SSI in hepato-pancreaticobiliary surgery patients with each independent variable.

**Results:** A total of 150 patients were included in the study. The mean age was 49.99 years, with females accounting for 62% of the participants. Ninety-six percent of surgeries were elective. Surgical site infections (SSIs) occurred in 27 patients (18.0%), the most common of whom presented with superficial incisional infections (14.0%). The average time to SSI onset was 6.9 days post-operatively. Multivariate logistic regression revealed that coagulopathy (AOR = 14.604; 95% CI: 1.068–199.769; p = 0.045), the presence of jaundice (AOR = 6.214; 95% CI: 2.180–17.714; p = 0.001), and postoperative hospital stay (AOR = 1.202; 95% CI: 1.058–1.366; p = 0.005) were independently associated with an increased risk of SSI.

**Conclusion:** The duration of stay in the hospital was a major factor in our study to impact the prevalence of surgical site infections; hence, it is important to carefully decide when to admit a patient and when to discharge.

## Introduction

Surgical site infections (SSIs) are infections that occur at the site of a surgical incision within 30 days of the procedure or within one year if an implant is involved(1,2). SSIs are potentially preventable complications following surgery and impose a significant burden in terms of patient morbidity, mortality, increased cost of treatment, and diminished quality of life for patients(2–4). SSIs can be classified into three categories: superficial incisional, deep incisional, and organ/space infections, each of which varies in severity and management(2).

The prevalence of SSIs in hepatopancreatobiliary surgery varies widely across different studies and settings. Research indicates that SSI rates can range from 0.64% to 54.7% depending on various factors, including the type of surgery performed, the patient population, and institutional practices(7,9–13). A systematic review of 25 articles on the incidence and risk factors for surgical site infections following hepatopancreatobiliary surgery from 2013--2022 reported rates of SSI ranging from 2%-54.7%, with an average of approximately 29.8%(14). The factors found by different studies to be associated with the development of SSIs in HPB surgery. Comorbidities such as diabetes mellitus, obesity, immunocompromised states and COPD were found to significantly increase the risk of SSIs. Patients with significant malnutrition also face heightened risks due to impaired wound healing and immune function(4,6,11,15,16).

The type of surgical procedure and its complexity play crucial roles in infection rates. Compared with laparoscopic approaches, open surgeries generally have higher SSI rates. Additionally, prolonged surgical duration has been correlated with increased infection risk(4,6,11,15–17). Adherence to sterile techniques, appropriate antibiotic prophylaxis, and the use of drains can influence the likelihood of SSIs. Studies emphasize the importance of administering prophylactic antibiotics within one hour before incision to reduce infection rates(4,16).

Effective wound care and monitoring for signs of infection are critical in the postoperative period. Patients who receive education on wound care and follow-up have been shown to have lower infection rates(2,4,16). In Ethiopia, various studies have explained the prevalence of SSIs and the factors associated with them in different healthcare settings. There appears to be a significant gap in the research specifically addressing the prevalence and risk factors associated with SSIs in the context of HPB surgery. Therefore, this study aimed to determine the prevalence and risk factors associated with SSIs in HPB surgery in selected hospitals in Addis Ababa.

Understanding the prevalence and factors associated with SSIs in patients undergoing HPB surgeries at Addis Ababa is of paramount importance. Identifying their prevalence and associated factors will enable targeted interventions to improve patient safety and outcomes. The findings of this study can help improve the overall quality of care for patients undergoing HPB procedures. These findings can support the development of hospital-wide and national guidelines and policies for the prevention and management of SSIs, particularly in HPB surgeries.

## Methods

### Study Setting and Period

A hospital-based prospective cross-sectional study was conducted in three major hospitals in Addis Ababa, Ethiopia: Tikur Anbessa Specialized Hospital, Yekatit 12 Specialized Hospital, and Menelik II Specialized Hospital. These hospitals are prominent healthcare facilities that provide hepatopancreaticobiliary(HPB) services via the HPB unit under the Department of Surgery of Addis Ababa University. Approximately 250–300 HPB surgeries, including complex hepatic and pancreatic resections, are performed yearly at Tikur Anbessa Hospital. A total of 300–400 HPB procedures, including cholecystectomies performed by general surgeons, are performed each year in the other two hospitals. These factors make the selected hospitals suitable sites for investigating the prevalence and factors associated with SSIs in HPB procedures.

The study took place over a period of 6 months. The study took place over a period of 6 months. It commenced on January 10, 2025 and concluded on June 30 2025. During this period, data, including preoperative assessments, intraoperative details, and postoperative follow-up data, were collected from patients undergoing HPB surgery to identify the occurrence of SSIs and associated risk factors.

### Study Design

An institution-based prospective cross-sectional study was conducted among hepato-pancreaticobiliary surgery patients in the selected hospitals Addis Ababa.

### Population

#### Source population

All patients who underwent hepatopancreaticobiliary surgeries at the selected hospitals in Addis Ababa were included.

#### Study population

All patients who met the inclusion criteria were hepatopancreaticobiliary surgery patients aged 18 years and above who had been managed surgically at the selected hospitals during the study period.

#### Data collection tools and techniques

The data were collected by medical doctors (general practitioners) via data collection questionnaires prepared by the research team. One surgical resident supervisor and one BSc nurse in each study setting were assigned for the data collection process, training was given to the data collectors by the principal investigator for two days on the objective of the study, how to approach participants, and how to fill and collect the relevant data via the data collection sheet. The Principal Investigator continuously supervised the data collectors. Sociodemographic and other patient-related factors were obtained directly from patients and patient medical charts. Data on the time of antibiotic prophylaxis administration and intraoperative doses were collected by direct observation and from patients’ operation notes. Data about antibiotics administered after the operation and the duration of administration were extracted from the patient medication chart and by direct observation. The patients were followed 30 days after the operation was performed for the status of their surgical site and overall condition. If there was any complaint, they were assigned to the operating hospital for confirmation of surgical site infection and further evaluation.

#### Sample size calculation

The sample size was calculated via the single population proportion formula and the double population formula via epi-info calc software.

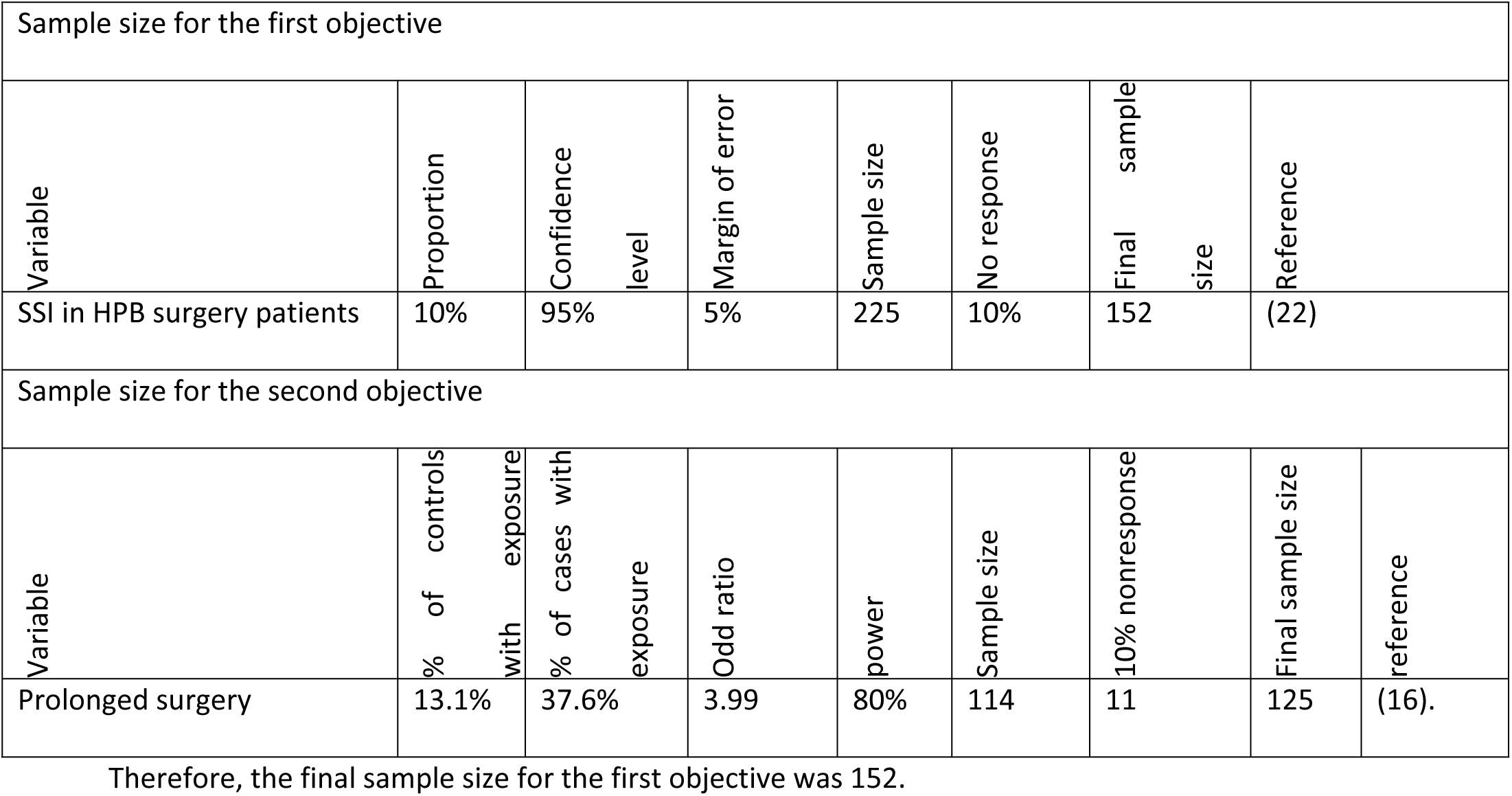

#### Sampling Procedure

All cases during the study period were selected until the calculated sample size was achieved during the study period.

#### Eligibility criteria

##### Inclusion criteria

Hepato-pancreaticobiliary surgery patients aged 18 years and above who were managed surgically at the selected healthcare facilities during the study period.

##### Exclusion criteria

✓ Multiple surgical procedures, including hepato-pancreaticobiliary surgery
✓ Patients who died in the OR table or before 24 hours post-surgery
✓ Laparoscopic cholecystectomy
✓ Patients lost to follow-up before 30 days
✓ Patient who died before 30 days without developing SSI

#### Study Variables

##### Dependent variable

✓ SSI in hepato-pancreaticobiliary surgery (yes/no)

##### Independent variables

➢ **Sociodemographic characteristics:** Age, Sex, Smoking, Alcohol, Residence
➢ **Health Status:** BMI, ASA Class
➢ **Comorbidity-related factors**: ASA class, Immobilization, Heart disease, Obesity, Diabetes Mellitus, Hypertension, CLD, COPD, Cardiovascular Diseases, HIV/AIDS, Coagulopathy, Malignancy, Alcoholic Liver Disease, Nonalcoholic Fatty Liver Disease, Previous infection
➢ **Surgical characteristics:** Elective vs Emergency Surgery, Types of Surgery/Procedures, Indications for surgery

➢ **Preoperative factors: Pr**eoperative hospital stay (days), Biliary malignancy, Liver cirrhosis Preoperative cholangitis, Preoperative biliary drainage, Presence of jaundice, Preoperative skin infection, Skin preparation type
➢ **Intraoperative:** Blood transfusion, Surgical type, Length of operation, Conversion to open surgery

➢ **Postoperative factors:** Prolonged hospital stays, External-internal drain, Pancreatic fistula, Postoperative biliary leakage, Postoperative anemia, Postoperative pneumonia
➢ **Preoperative Laboratory Parameters:** Hemoglobin, Albumin, WBC count, Platelet count, Direct bilirubin, Total bilirubin, ALT, AST, ALP, creatinine, BUN

➢ Postoperative Laboratory Parameters: Hemoglobin, WBC, Platelet count

#### Data Quality Control

The data collection sheet adopted from previous studies was checked for internal consistency and was capable of yielding the required data for the study, and some modifications were made. Training was given to the data collectors. The principal investigator also supervised the data collection process. Pretesting was performed in 5% of the questionnaires out of the study area by the principal investigator. The collected data were checked for completeness, consistency and clarity.

#### Operational definition

**Surgical site infection**: Incisional SSI is an infection that occurs within 30 days after the operation involving the skin, subcutaneous tissue, or deep soft tissue (e.g., fascia and muscle layers) at the incision site and at least one of the following:

1. Purulent drainage from the incision, with or without laboratory confirmation, from the incision
2. At least one of the following signs or symptoms of infection: pain or tenderness; localized swelling; redness; heat; or fever ≥38°C.
3. Spontaneous wound opening
4. An abscess or other evidence of infection involving the fascia or muscle layer found on direct examination, during reoperation, or by histopathological or radiological examination findings, culture of fluid or tissue in the organ/space;

**Multimorbidity**: living with two or more chronic illnesses

**Prolonged preoperative hospital stays:** Preoperative hospital stays greater than the 75th percentile of hospital stays (8, 23, 24).

**Prolonged surgery** was defined as surgery duration longer than the 75^th^ percentile of surgery duration (8, 23, 24).

**Prolonged postoperative hospital stays**- postoperative hospital stays greater than the 75^th^ percentile(25)

**Postoperative** anemia- hemoglobin level less than 10/dl after surgery(26)

**Postoperative pneumonia definition**: pneumonia occurring after 48–72 hours of hospitalization or surgery

**Massive/Major bleeding:** Bleeding greater than the 75^th^ percentile

#### Data entry and analysis

The data were checked for completeness entry by using Epi-data 4.1 exported, cleaned and analyzed via SPSS version 25. Descriptive statistics were run, and the generated data were compiled as proportions, frequency tables, charts, and graphs. Bivariate logistic regression analyses were performed to evaluate the associations of SSI in hepato-pancreaticobiliary surgery patients with each independent variable, and a p value<0.2 was used in the multivariable regression. To evaluate the strength of the association, both crude and adjusted odds ratios with 95% confidence intervals were calculated. The final model had a P value <0.05 and was considered statistically significant, and the strength of the association was determined from the adjusted odds ratio.

#### Ethical Considerations and Consent to Participate

The proposal was approved by the ethical committee of the Surgery Department College of Health Sciences of Addis Ababa University and the Addis Ababa Health Bureau for ethical approval under reference numbers A/A/T/2/039/17. An official letter of cooperation was written to the study hospitals to collect data. The study was conducted in accordance with the principles of the Declaration of Helsinki. All the study participants were informed about the purpose of the study, and written informed consent was obtained from study participants. The respondents had the right to refuse participation or terminate their involvement at any point during the interview. The information provided by each respondent was kept confidential. Furthermore, the reporting did not refer to a specific respondent.

## Results

A total of 150 patients who underwent hepato-pancreaticobiliary (HPB) surgeries across three tertiary hospitals, namely, Addis Ababa Tikur Anbessa Specialized Hospital (TASH), Yekatit 12 Hospital, and Minilik II Hospital, were included in this study.

### Sociodemographic characteristics of the patients

The mean age of the participants was 49.99 years (SD = 15.05), ranging from 19--80 years. The age group with the highest frequency was 36–50 years (37.3%). Females constituted 62.0% of the participants, whereas males accounted for 38.0%. Most patients (91.3%) had no history of smoking. In terms of alcohol consumption, 78.7% of the participants were nondrinkers, 18.0% were occasional drinkers, and 3.3% were regular drinkers (Table 1).

**Table 1.** Sociodemographic characteristics of the patients.

| Characteristic | Category | Frequency (%) |
| --- | --- | --- |
| Age | Mean | 49.99 years |
|  | Range | 19–80 years |
| Sex | Female | 62.0% |
|  | Male | 38.0% |
| Smoking Status | Nonsmoker | 91.3% |
|  | Former smoker | 6.7% |
|  | Current smoker | 2.0% |
| Alcohol Consumption | Nondrinker | 78.7% |
|  | Occasional drinker | 18.0% |
|  | Regular drinker | 3.3% |

The body mass index (BMI) classification revealed that the majority of participants (56.0%) had a normal weight (18.5–24.9 kg/m²). A total of 30.7% were underweight, with a BMI of less than 18.5 kg/m². Moreover, 13.3% of the participants were categorized as overweight (25–29.9 kg/m²).

### Comorbidities

Diabetes mellitus and hypertension were the most common comorbidities as shown in Table 2.

**Table 2.** Comorbidities and physical status of the patients.

| **Comorbidity** | **Frequency (n)** | **Percentage (%)** |
| --- | --- | --- |
| Diabetes Mellitus | 18 | 12.0% |
| Hypertension | 35 | 23.3% |
| Chronic Liver Disease (CLD) | 4 | 2.7% |
| Cardiovascular Disease | 2 | 1.3% |
| Chronic Obstructive Pulmonary Disease (COPD) | 2 | 1.3% |
| HIV/AIDS | 7 | 4.7% |
| Coagulopathy (Bleeding Disorders) | 3 | 2.0% |
| Obesity | 1 | 0.7% |
| Alcoholic Liver Disease | 0 | 0.0% |
| Nonalcoholic Fatty Liver Disease | 0 | 0.0% |
| Immobilization | 0 | 0.0% |
| **ASA Physical Status Classification** |  |  |
| ASA I | 19 | 12.7% |
| ASA II | 108 | 72.0% |
| ASA III | 23 | 15.3% |
| **BMI Classification** |  |  |
| Underweight (<18.5 kg/m<sup>2</sup>) | 46 | 30.7% |
| Normal weight (18.5–24.9 kg/m<sup>2</sup>) | 84 | 56.3% |
| Overweight (25–29.9 kg/m<sup>2</sup>) | 20 | 13.3% |

On the basis of the American Society of Anesthesiologists (ASA) physical status classification, the majority of patients were classified as ASA II, accounting for 72.0% of the study population. This was followed by ASA III, comprising 15.3%, and ASA I, representing 12.7% of the participants.

### Surgical characteristics of patients

Nearly all surgeries were elective (96.0%), with six emergency cases (4.0%). The most commonly performed procedure among the study participants was open cholecystectomy, accounting for 36.7% of the cases, followed by bypass surgery at 16.0%. The most common indications for surgery were cholelithiasis and periampullary carcinoma (Table 3).

**Table 3.** Indications for HPB surgery in patients.

| **Indication** | **Frequency (n)** | **Percentage (%)** |
| --- | --- | --- |
| Cholelithiasis | 47 | 31.3% |
| Periampullary Carcinoma | 27 | 18.0% |
| Choledocholithiasis | 12 | 8.0% |
| Hepatocellular Carcinoma (HCC) | 10 | 6.7% |
| Cholangiocarcinoma | 3 | 2.1% |

### Preoperative and Intraoperative Factors

The mean duration of the preoperative hospital stay was 4.81 days (SD = 4.03), ranging from 1--45 days. The majority of patients stayed for more than 3 days prior to surgery. Among the study participants, liver cirrhosis was identified in 2.7% of them. Preoperative cholangitis was observed in 20.0% of patients, and 6.7% underwent preoperative biliary drainage. Jaundice was present in 33.3**%** (n = 50) of the patients. Whole blood counts and liver enzyme and bilirubin levels were collected from the patients preoperatively (Table 4).

**Table 4.** Preoperative laboratory values of the patients.

| Parameter | Category | Frequency (n) | Percentage (%) |
| --- | --- | --- | --- |
| **Hemoglobin (g/dL)** | ≥11 | 121 | 80.7 |
|  | 7–10.9 | 29 | 19.3 |
| **WBC Count (×10<sup>3</sup>/μL)** | <4 x10<sup>3</sup> | 13 | 8.7 |
|  | 4–12 x10<sup>3</sup> | 115 | 76.7 |
|  | >12 x10<sup>3</sup> | 22 | 14.7 |
| **Platelet Count (×10<sup>3</sup>/μL)** | ≥150 | 131 | 87.4 |
|  | 100–149 | 14 | 9.3 |
|  | <100 | 5 | 3.3 |
| **Serum Albumin (g/dL)** | ≥3.5 | 65 | 50.0 |
|  | 2.5–3.4 | 43 | 33.3 |
|  | <2.5 | 5 | 4.0 |
|  | Not available | 37 | 24.7 |
| **Direct Bilirubin (mg/dL)** | <3 | 86 | 63.4 |
|  | 3–8 | 21 | 15.2 |
|  | 8–15 | 15 | 11.0 |
|  | >15 | 6 | 4.1 |
| **Total Bilirubin (mg/dL)** | <5 | 105 | 71.2 |
|  | 5–10 | 25 | 17.0 |
|  | 10–20 | 13 | 8.8 |
|  | >20 | 4 | 2.7 |
| **ALT Level** | Normal | 99 | 66.0 |
|  | 2× ULN | 49 | 32.7 |
|  | ≥3× ULN | 2 | 1.3 |
| **AST Level** | Normal | 111 | 74.0 |
|  | 2× ULN | 34 | 22.7 |
|  | ≥3× ULN | 5 | 3.3 |

**Table 5.** Bivariate logistic regression between variables and SSI.

| Variable | COR | 95 % CI | p-value |
| --- | --- | --- | --- |
| Age | 1.02 | 0.99 – 1.05 | 0.109* |
| Sex (Female vs Male) | 0.72 | 0.31 – 1.68 | 0.447 |
| Residential Area (Urban vs Rural) | 1.23 | 0.46 – 3.32 | 0.682 |
| Smoking Status | 2.02 | 0.49 – 8.37 | 0.333 |
| Alcohol Consumption | 1.71 | 0.64 – 4.60 | 0.284 |
| Body Mass Index (BMI) | 0.90 | 0.57 – 1.43 | 0.663 |
| Diabetes Mellitus | 2.02 | 0.65 – 6.25 | 0.225 |
| Hypertension | 0.71 | 0.25 – 2.02 | 0.515 |
| Coagulopathy/Bleeding Disorders | 9.76 | 0.85 – 111.83 | 0.067* |
| ASA Class |  |  |  |
| ASA I vs III | 0.24 | 0.06 – 0.97 | 0.044* |
| ASA II vs III | 0.10 | 0.04 – 0.29 | <0.001* |
| Type of Surgery | 0.91 | 0.10 – 8.10 | 0.931 |
| Duration of Preoperative Stay | 1.145 | 0.987-1.329 | 0.073* |
| Liver Cirrhosis | 1.54 | 0.15 – 15.38 | 0.714 |
| Cholangitis | 1.93 | 0.75 – 4.98 | 0.172* |
| Preoperative Biliary Drainage | 0.49 | 0.06 – 4.02 | 0.504 |
| Albumin | 0.95 | 0.49 – 1.84 | 0.886 |
| Preoperative Direct Bilirubin | 0.34 | 0.14 – 0.80 | 0.014* |
| Preoperative Total Bilirubin | 0.44 | 0.20 – 0.94 | 0.033* |
| Preoperative ALT | 0.38 | 0.14 – 1.05 | 0.063* |
| Preoperative AST | 0.70 | 0.29 – 1.71 | 0.438 |
| Preoperative ALP | 0.78 | 0.45 – 1.35 | 0.381 |
| Preoperative Creatinine | 0.64 | 0.08 – 5.41 | 0.680 |
| Preoperative Skin Infection | 1.52 | 0.58 – 4.03 | 0.398 |
| Duration of surgery | 1.001 | 0.996 - 1.006 | 0.669 |
| Postoperative Duration of Stay | 1.21 | 1.07 – 1.38 | 0.003* |
| ICU Admission | 2.38 | 0.41 – 13.71 | 0.332 |
| Postoperative Antibiotics | 0.98 | 0.39 – 2.45 | 0.970 |
| Use of Postoperative Drains | 1.74 | 0.75 – 4.05 | 0.199* |
| Postoperative Hemoglobin | 0.94 | 0.76 – 1.16 | 0.540 |
| Postoperative WBC | 1.00 | 1.00 – 1.00 | 0.614 |
| Postoperative Platelet Count | 1.00 | 1.00 – 1.00 | 0.559 |

Antibiotic prophylaxis was administered to all patients in the study, predominantly with ceftriaxone (42.7%) and combinations with metronidazole. Intraoperative spillage occurred in one patient (0.7%). Preoperative skin preparation predominantly involved the use of iodine-based solutions, which were applied in 96.0% of patients. Alcohol-based solutions were used in 35.3% of the cases. Additionally, preoperative infections were identified in 20.0% of the study participants.

Among the 150 patients studied, conversion from laparoscopic to open surgery was required in only 2 patients (1.3%). The mean operative time was 171.2 minutes (SD ±78.4), with a range of 60--480 minutes. The most frequently recorded duration was between 60 and 120 minutes. The average estimated blood loss (EBL) was 266 ml (SD ±264.0). Intraoperative blood transfusions were administered to 7.3% of the patients.

### Postoperative factors

The mean postoperative hospital stay was 4.49 days (SD ±3.28), with a median stay of 4 days and a range of 1--25 days, with the most common stay being 3 days. Postoperative ICU admission was required for 6 patients (4.0%), whereas 144 patients (96.0%) were managed without intensive care. Postoperative antibiotic therapy was administered to the majority of patients (70.7%, n = 106), whereas 44 patients (29.3%) did not receive antibiotics after surgery. Surgical drains were placed in 72 patients (48.0%), whereas in 78 patients (52.0%), drains were not inserted. Postoperative infections, excluding surgical site infections (SSIs), are relatively uncommon. Pneumonia occurred in 8 patients (5.3%), urinary tract infections (UTIs) occurred in 3 patients (2.0%), and other infections, such as ascending cholangitis or seroma with systemic symptoms, were reported in 1 patient each.

Reoperations were required in 4 patients (2.7%), primarily because of abdominal collection (n=1), postoperative collection (n=1), or biliary leakage (n = 1). Postoperative blood transfusions were administered to 8 patients (5.3%). No cases of postoperative pancreatic leakage were reported in the study population. A prolonged total duration of stay was found in 39 (26.0%) patients who stayed in the hospital for 6 days preoperatively, as determined by the 75^th^ percentile of the duration of stay for all patients.

### Surgical Site Infections (SSI)

Among the 150 patients included in the study, 27 (18.0%) developed surgical site infections (SSIs), whereas the remaining 123 patients (82%) presented no signs of infection, as shown in Figure 1. Among the 27 patients who developed SSI (18% of the total patients), the mean time to onset was 6.3 days post-surgery, with a median of 6 days post-surgery. The minimum time to onset was 2 days, and the maximum was 15 days. The second most frequently reported onset day was the postoperative day.

**Figure 1.**
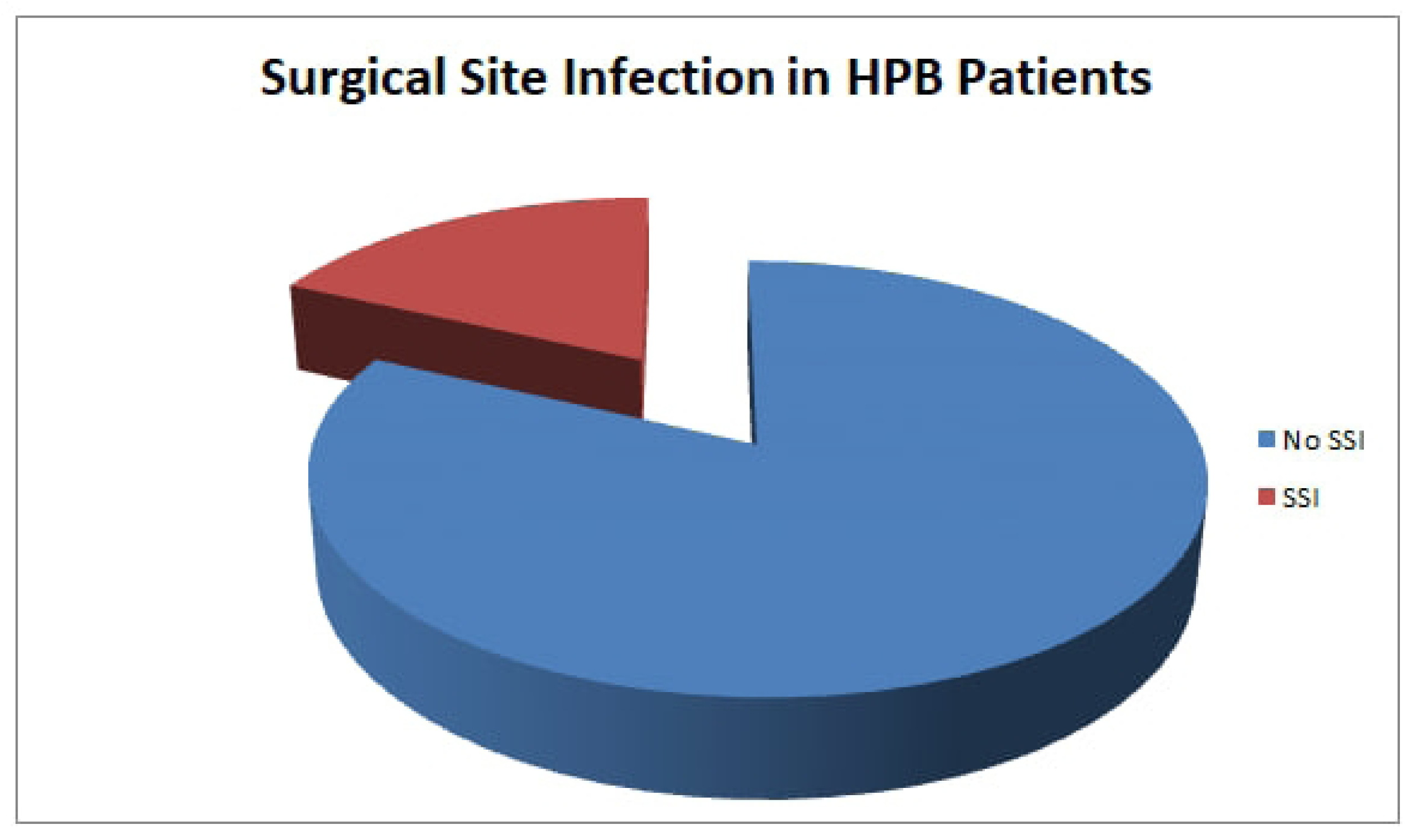
Proportion of patients with hepatopancreaticobiliary diseases having surgical site infection.

Among those who developed SSI, various wound characteristics were observed. Redness or erythema was noted in 13 patients (8.7%), purulent discharge in 19 patients (12.7%), wound dehiscence in 2 patients (1.3%), and intra-abdominal collections in 5 patients (3.3%). The mean time to onset of SSI was 6.93 days (SD ±3.1), with the earliest case occurring on postoperative day 2 and the latest occurring on day 15. The majority of SSIs were diagnosed between the second and ninth postoperative days. Among the 27 patients who developed surgical site infections (SSIs), the majority were classified as superficial incisional infections, accounting for 21 cases (14.0%). Deep incisional SSIs were identified in 3 patients (2.0%), and organ/space infections were also observed in 3 patients (2.0%). Microbiological cultures were performed in a limited number of cases. Among the 150 patients, 22 (14.7%) had no cultures taken, 4 (2.7%) had unspecified microbiological results, 1 (0.7%) had a fungal infection, and another 1 (0.7%) had a polymicrobial infection.

With respect to the management of SSIs, 12 patients (8.0%) were treated with antibiotics alone, whereas 21 patients (14.0%) received wound dressing changes. Two (1.3%) of the patients underwent surgical debridement as a standalone treatment. Readmission occurred in 3 patients (2.0%) who required further management for surgical site infections. Mortality was reported in one patient (0.7%), with the primary cause of death identified as septic shock.

### Factors associated with SSI

To identify factors associated with surgical site infections (SSIs), bivariate logistic regression analysis was performed. A p value threshold of 0.2 was used to screen variables for inclusion in further analysis. The following variables were assessed individually against the presence of SSI. Age, ASA class, coagulopathy, duration of preoperative stay, cholangitis, direct bilirubin, total bilirubin, ALT, presence of jaundice, postoperative hospital stay and drainage tube use were factors that significantly affected the results of the bivariate logistic regression.

Variables that were found to be significant via bivariate logistic regression were further entered into multivariate logistic regression. Statistically significant associations were found between SSI and ASA class II, coagulopathy, jaundice, total bilirubin and postoperative stay. Coagulopathy was strongly associated with SSI, as patients with coagulopathy were fourteen times more likely to develop SSI. A lower total bilirubin level was associated with a reduced risk of SSI. Patients who had jaundice were also six times more likely to develop SSI (AOR = 6.214), as shown in Table 7.

**Table 7.** Multivariate logistic regression of variables.

| Variable | AOR | 95% CI | p value |
| --- | --- | --- | --- |
| ASA Class II | 0.110 | 0.039 – 0.312 | <0.001* |
| ASA Class I | 0.303 | 0.073 – 1.248 | 0.098 |
| Coagulopathy | 14.604 | 1.068 – 199.769 | 0.045* |
| HIV | 4.668 | 0.826 – 26.400 | 0.081 |
| Age | 1.016 | 0.986 – 1.046 | 0.299 |
| Preoperative Stay | 1.129 | 0.975 - 1.308 | 0.104 |
| Jaundice | 6.214 | 2.180 – 17.714 | 0.001* |
| Total Bilirubin | 0.424 | 0.190 – 0.948 | 0.037* |
| Direct Bilirubin | 0.891 | 0.790 – 1.005 | 0.061 |
| Postoperative Stay | 1.202 | 1.058 – 1.366 | 0.005* |
| ALT | 0.460 | 0.152 – 1.395 | 0.170 |
| Drainage Tube Use | 1.486 | 0.591 – 3.739 | 0.400 |

## Discussion

This study revealed a surgical site infection (SSI) rate of 18.0% following hepato-pancreaticobiliary surgeries across three tertiary hospitals in Addis Ababa, Ethiopia. This prevalence falls within the range reported globally, ranging from 0.64% to 54.7%, depending on the type of surgery, patient profile, and institutional practices (18,19,20,21,22). This finding aligns closely with the 25.1**%** SSI rate reported in a German study(23), the 17.8% rate reported in Japan (9), and the 32% rate reported following pancreaticoduodenectomy in Thailand(24). However, this incidence is slightly higher than the 14.1**%** pooled incidence reported by the WHO for HPB procedures in low- and middle-income countries(25). The majority of SSIs in this study were superficial incisional infections (14.0%), which is consistent with the findings of previous studies (20,23,26), where superficial infections often account for the largest proportion of SSIs after HPB surgery. Overall, the high prevalence of SSI is high across countries, which warrants that all stakeholders reduce its prevalence.

Unlike findings from multiple studies where male sex, age above 50 **years**, and high BMI were significantly associated with SSI(19,24), our current study revealed no statistically significant associations with age, sex, or BMI. In this study, HIV/AIDS was not associated with SSI (p = 0.081). In contrast, immune-compromised patients, particularly those with HIV, are at increased risk of SSI due to impaired immune responses and poor tissue healing(11,22,26,27). Another study performed by David K. Warren et al. highlighted immune dysfunction and malnutrition as important risk factors for infection after abdominal surgery(11). Interestingly, diabetes mellitus, another commonly cited risk factor (11,22,27), was not statistically significant in this study, which contrasts with multiple studies identifying it as an independent predictor of SSI. This difference may be due to the smaller diabetic population in our study and the overall limited sample size.

Preoperative laboratory parameters such as elevated total bilirubin (AOR =0.424, 95% CI: 0.**190–0**.948) **were** significant in multivariate analysis, where low levels of total bilirubin were protective against SSI. However, BUN levels were not associated with SSI in this study. Prior studies have identified elevated direct bilirubin (>15 mg/dL) as a risk factor for SSI(22), whereas the association between renal dysfunction (reflected in BUN) and SSI has been less consistently reported. In contrast, the duration of surgery, although a significant factor in other studies (22,27), was not associated with SSI in our study (p = 0.669). This might be explained by the fact that surgical time data were not available for all patients, reducing the power to detect an effect.

Postoperative hospital stay was another strong independent predictor (AOR = 1.202, p = 0.005), aligning with studies from Ethiopia and abroad that associated prolonged hospitalization with a greater risk of hospital-acquired infections (22,26,28,29). The effect of prolonged postoperative stay may reflect early complications in two ways and may also act as a source of further nosocomial exposure. Postoperative complications such as anastomotic leakage, bile leakage, and the need for reoperation were not statistically significant in our study. This contradicts robust evidence from Suragul et al., who reported a strong association between postoperative bile leakage and SSI (OR = 2.78) (24), and from Sugiura et al., who reported that postoperative pancreatic fistula, drain use, and prolonged drainage were key risk factors(27). Our inability to detect these associations could stem from the small sample size; for example, there were only three cases of anastomotic leakage and two cases of bile leakage in our study, limiting statistical power. Other factors, such as intensive care unit (ICU) admission, surgical **drain use**, and postoperative antibiotic use, were not associated with SSI in our study. These findings contrast with reports suggesting that excessive drain use or inadequate prophylaxis may increase risk (22,27).

In our setup, ceftriaxone with metronidazole was the most commonly used prophylactic antibiotic, which may be insufficient for high-risk HPB surgeries compared with regimens used in Japanese and German centers (22,26). The culture results were recorded in only a few cases, and polymicrobial or fungal infections were rare. Most patients were managed with wound care and antibiotics. One patient (0.7%) died from septic shock, emphasizing the potential severity of SSIs in HPB surgery and reinforcing the need for prevention and timely management.

### Conclusions and recommendations

The prevalence of surgical site infections remains high. Efforts to prevent SSI should be given more priority and investment. The duration of stay in the hospital was a major factor in our study to impact the prevalence of surgical site infections; hence, it is important to carefully decide when to admit a patient as well as when to be discharged. A high index of suspicion should be given to immunocompromised individuals for both early detection and prevention of surgical site infections.

### Limitations

Although a prospective cross-sectional study design was used, our ability to identify causal relationships between identified risk factors and surgical site infections is limited.

## List of abbreviations

AOR: adjusted odds ratio
ASA: American Society of Anesthesiology
CDC: Control Disease Prevention
HAIs: Healthcare-associated infections
HPB: Hepatopancreatobiliary
OR: Odd ratio
SSI: Surgical site infection
TASH: Tikur Anbessa Specialized Hospital

## Availability of data and materials

The datasets used in this study are available from the corresponding author upon request.

## Authors’ contributions

GE wrote the proposal, ST collected the data and AM analyzed the data. AM and GE wrote the final manuscript. SW reviewed and provided directions during the write-up process.

## Declaration of Generative AI and AI-assisted technologies in the writing process

During the preparation of this work, the author used Curie to improve the language. After using this tool/service, the author reviewed and edited the content as needed and takes full responsibility for the content of the published article.

## Disclaimer

The views and opinions expressed in this article are those of the author/authors and are the product of professional research. They do not necessarily reflect the official policy or position of any affiliated institution, funder, agency, or publisher. The authors are responsible for this article’s results, findings, and content.

